# Horizon, closing the gap between cinematic visualization and medical imaging

**DOI:** 10.1101/2023.07.11.23292519

**Authors:** Javier Guaje, Serge Koudoro, Eleftherios Garyfallidis

## Abstract

Medical imaging has become a fascinating field with detailed visualizations of the body’s internal environments. Although the field has grown fast and is sensitive to new technologies, it does not use the latest rendering techniques available in other domains, such as day-to-day movie production or game development. In this work, we bring forward Horizon, a new engine that provides cinematic rendering capabilities in real-time for quality controlling medical data. In addition, Horizon is provided as free, open-source software to be used as a foundation stone for building the next generation of medical imaging applications. In this introductory paper, we focus on the extensive development of advanced shaders, which can be used to highlight untapped features of the data and allow fast interaction with machine learning algorithms. In addition, Horizon provides physically-based rendering capabilities, the epitome of advanced visualization, adapted for the needs of medical imaging analysis practices.

## 1. Introduction

Scientific data is continuously increasing in size and complexity, and so is the necessity for efficient and reliable visualization tools. Due to this need, computing has become a fundamental part of the modern scientific method. Consequently, visualization has become indispensable for interpreting the acquired, processed, and generated data. Some authors have defined *visualization* as the process of providing new scientific insight through visual methods (Hansen and Johnson, 2011). Furthermore, visualization promotes new forms of analysis, assessment of theoretical models, debugging and fine-tuning computational code, and science communication to broader audiences. *Scientific visualization* is the research field that studies all these use cases and mainly all the processes that transform data into rendered images (Wald et al., 2016). These processes follow the next pipeline: data analysis, filtering, mapping, and rendering. Although a big part of this approach focuses on data analysis, the last stage of the pipeline is rendering, in which data represented as a set of 3D geometric or volumetric primitives is transformed into 2D frames.

In contrast to production rendering engines, real-time rendering engines synchronously produce images, i.e., the renderer shows an image, and the viewer might act or react to it, providing feedback to an application that informs the rendering engine to generate the following appropriate image. To ensure that both the application and the rendering engine satisfy real-time quality, they must respond quickly to the user’s input so the viewer does not perceive the individual images but instead experiences immersion in a dynamic environment (Akenine-Moller et al., 2019). Given the growing demand for more realistic video games and the response from GPU (Graphics Processing Units) manufacturing companies, these engines have achieved significant progress, producing impressive graphics when the appropriate algorithms are used. These include physically-based rendering (Shirley, 1991), hard and soft shadowing (Crow, 1977; Williams, 1978), ambient occlusion (Miller, 1994), and ray tracing (Kajiya, 1986; Hofmann, 1990; Whitted, 2005). Although many of these techniques were initially production rendering techniques, incremental GPU improvements have made interactive use cases possible. The necessities of real-time scientific visualization are not far from the ones of the gaming industry. Some initiatives have taken rendering techniques commonly seen in movie making and game development to offer them to the scientific community (Stone et al., 1998; Wald et al., 2016; Garyfallidis et al., 2021).

Driven by a different motivation, production rendering seeks the highest visual quality at the expense of interactivity. Since the majority of these visual products are choreographed, it is common to use computing-demanding techniques, such as but not limited to, variations of ray tracing algorithms (Kajiya, 1986; Hofmann, 1990; Whitted, 2005). Although this type of rendering is typically used in movie production and photorealistic rendering for design and prototyping, it can also be used for scientific purposes (Stone et al., 1998; Wald et al., 2016; Garyfallidis et al., 2021). Cinematic scientific visualization is a recent application of production rendering whose purpose is not data exploration or analysis but the communication of scientific concepts (Borkiewicz et al., 2020). This application presents scientific data more clearly, aesthetically, and pleasantly than traditional visualization.

Medical imaging as a scientific domain is a very active and thriving field. In consequence, many standards in the area are still being proposed. A particular case is the multiple file formats created as a pursued goal or a byproduct of research. Its adoption by the scientific community is given by its accessibility and compatibility with existing and well-established tools and frameworks. However, supporting such a collection of file formats is challenging, and only a few projects exist that are doing this and open-sourcing it. Furthermore, the panorama is equally challenging when finding visual representations for such formats. Besides, modern generic medical visualization engines are not open-source or can not be easily extended.

In the medical imaging field, for example, most well-known and established analysis frameworks already include a visualization solution. Some projects in this category are FSLEyes in FSL (McCarthy, 2022), Mrview included in Mrtrix (Tournier et al., 2019), and Freeview part of FreeSurfer (Fischl, 2012). Nevertheless, the focus of these frameworks lies in the included analysis methods. In another category, DSI Studio (Yeh et al., 2010) and TrackVis (Wang et al., 2007) focus primarily on tractography analysis. Finally, there are tools with extensive visualization options such as MITK (Wolf et al., 2004), 3D Slicer (Kikinis et al., 2014) and MRIcroGL (Rorden and Brett, 2000). However, none of these tools provide real-time physically-based rendering capabilities. In that line of research, only cinematic rendering (Eid et al., 2017) has approached the techniques used in photorealistic rendering and used them to get more accurate 3D reconstructions from computerized tomography (CT) and magnetic resonance images (MRI). This work showed how more realistic representations of light interaction improved the spatial perception of objects in the scene.

This work introduces a novel open-source tool capable of visualizing many medical and commonly used file formats (NIfTI (NIfTI Documentation), TRX (Rheault et al., 2022), TRK (Wang et al., 2007), TCK (Tournier et al., 2012), GIfTI (Harwell et al., 2008), among others). In addition, it presents new techniques and ideas to explore and analyze medical images interactively, as well as state-of-the-art shading algorithms. These methods were added so scientists with no programming skills could use them. This proposed tool is called Horizon and comes integrated into one of the most popular frameworks to conduct diffusion magnetic resonance imaging (dMRI) analyses, DIPY (Diffusion Imaging in PYthon) (Garyfallidis et al., 2014). This new tool provides interactive and real-time cinematic rendering for medical imaging data. Therefore, Horizon interfaces advanced rendering technologies and medical imaging technologies. This is why this name was used, inspired by the fact that the actual horizon separates the sky (in our case, advanced rendering technologies) from the earth (in our case, medical imaging technologies). Horizon is partially written in Python and partially written in Shading Languages (primarily GLSL). Python is widely used in the scientific community and does not require compilation which facilitates the distribution of scripts and libraries. GLSL is also compiled automatically by the GPU. Additionally, Horizon can be used interactively in Jupyter-based environments, which makes it perfect for exploratory analyses. For graphics acceleration, Horizon includes elements written in GLSL, which is the primary shading language for OpenGL. These elements are executed in parallel by the GPU and are used for tasks like geometry amplification and lighting. This new addition follows both DIPY’s (Garyfallidis et al., 2014) and FURY’s (Garyfallidis et al., 2021) open principles, which means the design of the tool is modular and documented in a way such that researchers can see the underlying details. We expect researchers to find these features useful and take advantage of the new shading capabilities to improve the communication of their findings and extend them as their research practices require.

## 2. Methods

### 2.1. Cinematic scientific visualization

Horizon includes a rendering engine that supports Physically Based Rendering (PBR) (Shirley, 1991) capabilities. PBR engines aim to simulate light properties when interacting with objects in the scene in a physically plausible way. In order to achieve this, light is studied and simulated from a conductive point of view (Hoffman, 2013). In other words, light interacts differently with distinct objects’ materials. In PBR, materials are divided into two categories based on their conductive properties: dielectrics and metals (Shirley, 1991). *Metallic* is the first parameter of Horizon’s PBR engine. It takes values between 0 and 1, where the former represents dielectric materials and the latter metallic materials. Although, in principle, this parameter should be binary for only deciding the conductive properties of the surface, having intermediate values between 0 and 1 provides the flexibility for simulating more complex processes such as dust on metallic surfaces.

In order to be considered physically based, a rendering engine also must satisfy the following conditions: 1) It should be based on a microfacet surface model, 2) It should be energy conserving, and 3) It should use a physically based Bidirectional Distribution Function (BDF) (Hoffman, 2013).

A microfacet surface model states that any surface at a microscopic scale is composed of infinitesimal fully reflective mirrors called microfacets (Torrance and Sparrow, 1967). These mirrors can be almost perfectly or randomly aligned along the surface of an object (see midsection of Fig. 1). More chaotic alignment of these microfacets produces rougher surfaces which tend to scatter the incoming light rays in different directions. Conversely, less rough surfaces are more likely to reflect light in the same direction. This interaction constitutes the second parameter of Horizon’s rendering engine. The *roughness* parameter takes a value between 0 and 1, which is used to calculate the portion of microfacets aligned to the *halfway vector*. This vector represents the bisecting vector between the incoming *light* vector (***l***) and the outgoing *view* vector (***v***) on a surface’s point (see midsection of Fig. 1). It can be calculated as follows:

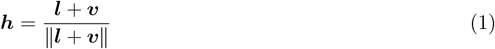

**Figure 1.**
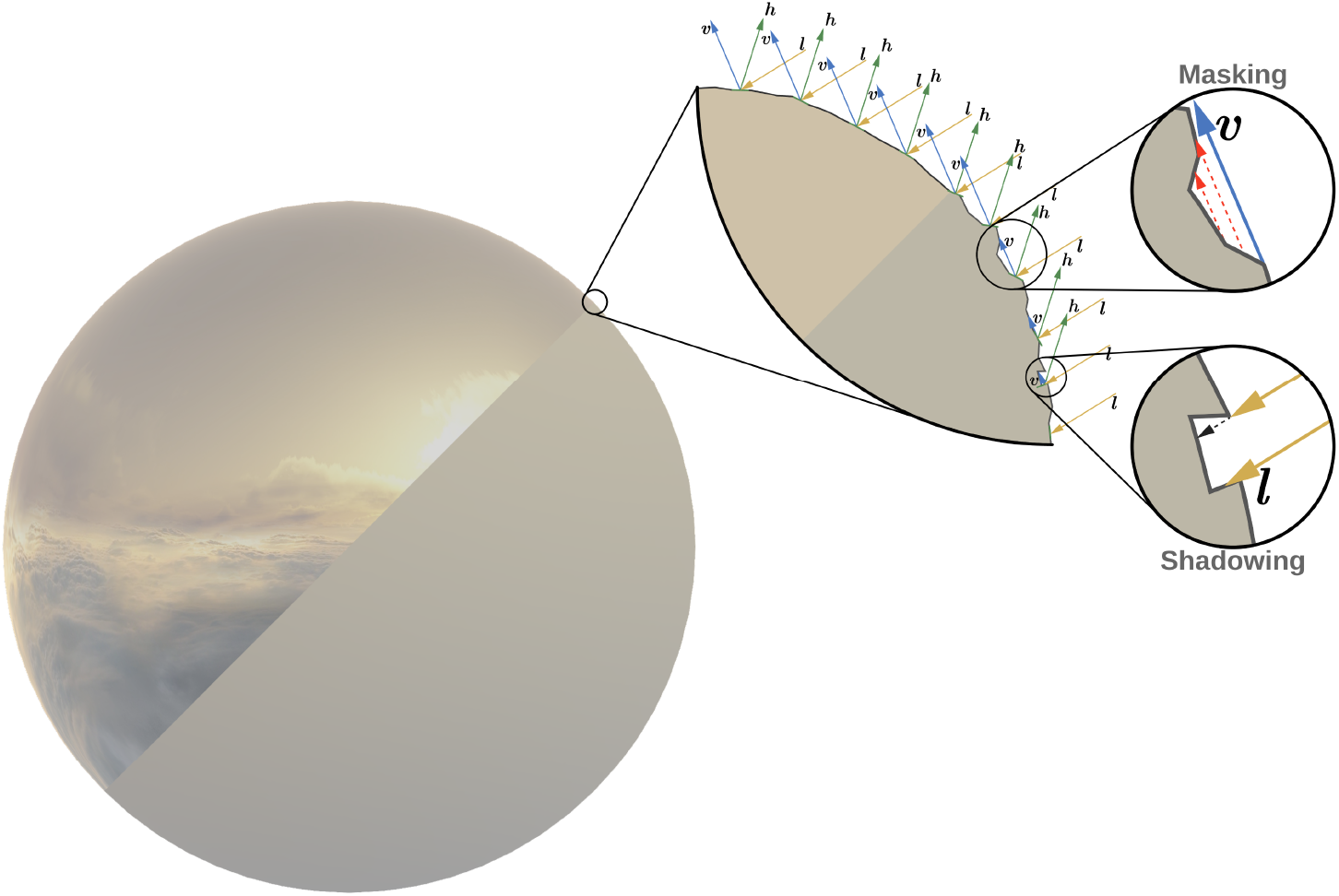
Diagram of the distribution of microfacets on smooth and rough surfaces. The sphere on the **left** simulates two physical properties. On **top** is a smooth surface, and on the **bottom** is a rough surface. The first zoomed-in section (**center**) illustrates how microfacets might align on such surfaces. The **rightmost** section shows two different occlusion effects present on rough surfaces.

**Figure 2.**
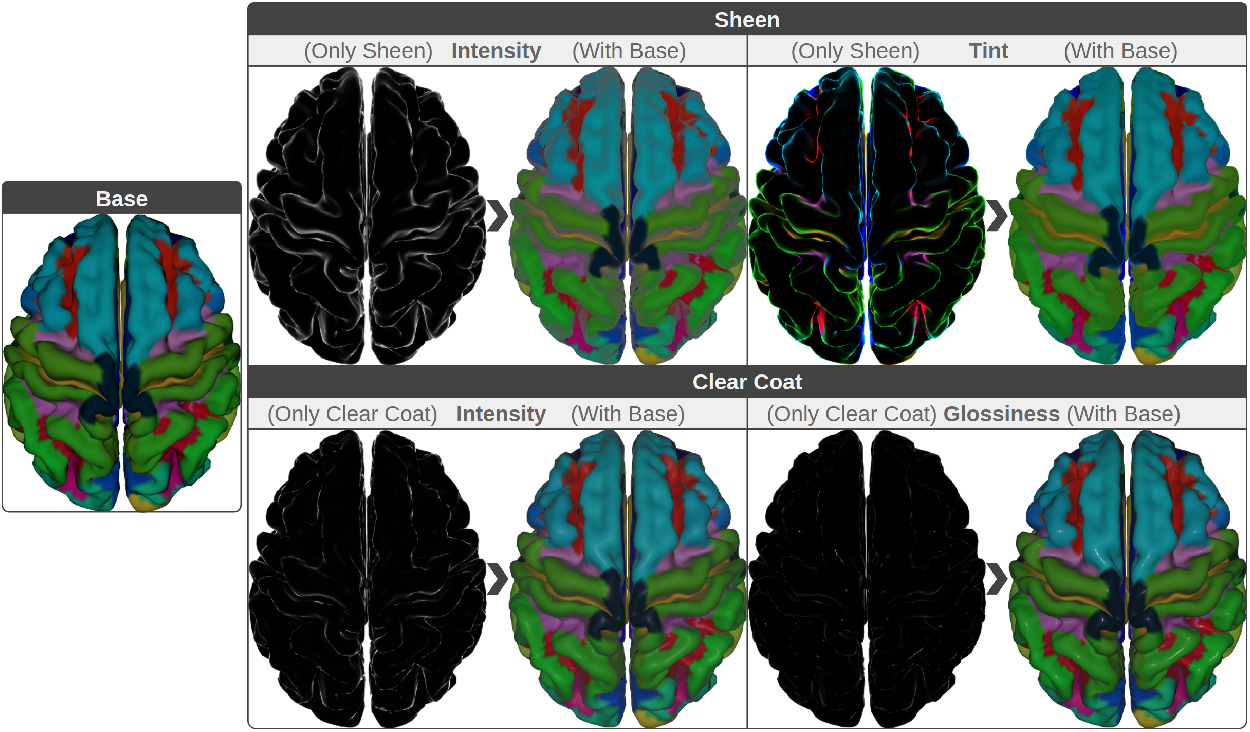
Demonstration of *Sheen* and *Clear Coat* effects. On the **left**, we see the base PBR colors. On the **right**, we see how *Sheen* (**top**) and *Clear Coat* (**bottom**) will change the base color. To clarify the advantage, we can see the effects isolated, assuming the base colors are zero (black) or together with the actual base colors (colored). Notice how some effects, such as *Clear Coat Glossiness*, have a smaller magnitude and others, such as *Sheen Tint*, have a more profound effect. When combined with the base color, such effects can provide more information about the underlying geometry of the surface.

PBR engines should also follow the energy conservation principle; this means the outgoing light energy should not exceed the amount of incoming light energy. Here, it is also essential to highlight the differences between *diffuse* and *specular* light. When a light beam hits a point on a surface, it is divided into a *refracted*, and a *reflected* part. Refracted light is the portion absorbed by the surface, known as diffuse lighting. Reflected light, on the other hand, is the part that gets directly mirrored instead of entering the surface, also called specular lighting.

The last element of the PBR engine is the BDF, which is a probability density function that takes the incoming *light* direction (***l***) and an outgoing *view* direction (***v***) for each point on the surface. BDFs have different types depending on the specific behavior of light that is desired to model. The most common and basic are the ones based on modeling the reflectance of different materials, also known as Bidirectional Reflectance Distribution Functions (BRDFs). The supported BDF in Horizon is a microfacet BRDF with custom diffuse component (*f*_*d*_) and a specular component based on the *Cook-Torrance* model (*f*_*s*_). Horizon’s BRDF is inspired by Disney’s “principled” shading approach and is represented with the following expression:

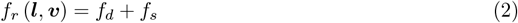

There are several methods to model the diffuse component of the BRDF, some more realistic than others. The counterpart of these methods is that they tend to be computationally more expensive, and in most cases, the *Lambertian* approach is sufficient enough. This method is described as:

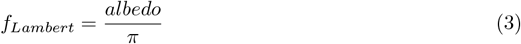

*albedo* is the surface color at that point and is divided by *π* to normalize the diffuse light. Though simple and functional, a major caveat of this method is that it tends to be darker around the edges. A way to address this issue and make it more physically plausible is by adding a *Fresnel* factor. This factor accounts for the portion of the light that gets reflected and refracted depending on the viewing angle. Although the *Fresnel* equation is complex to be used in many applications, the *Schlick* approximation is sufficient for our purposes and is given by:

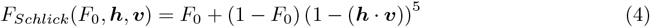

Where *F*_0_ is the base reflectivity and is constant for every material. From this expression it is possible to calculate the *Fresnel* factor, which denoted by (1 *−* (***l*** *·* ***n***)) (1 *−* (***n*** *·****v***)), where ***n*** represents the surface’s normal vector. Then a variation of this factor will be added to the diffuse term to account for darker effects. In particular, the approach followed introduces a transition between a *Fresnel* shadow for smooth surfaces and an added highlight for rough surfaces. Putting all together, the final diffuse component included in Horizon is:

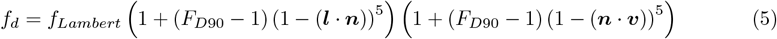

*F*_*D*90_ is a customized diffuse *Fresnel* factor that tweaks the glancing retroreflection response based on the surface’s *roughness*. It is calculated using the following equation:

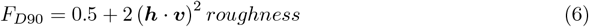

This produces a *Fresnel* shadow that reduces the incident reflectance by half at sharp angles for smooth surfaces and increases the response by up to 2.5 for *rough* surfaces.

*Subsurface* is an additive parameter that allows blending between the previously described diffuse component and one inspired by Hanrahan-Krueger subsurface BRDF (**?**). This is useful for giving a subsurface appearance on distant objects and on objects where the average scattering path length is small; it is not a substitute for an entire subsurface transport as it will not bleed light into the shadows or through the surface.

The second major term of Horizon’s BRDF (Eq. 2) is the specular reflection (*f*_*s*_) which is given by:

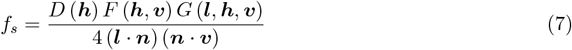

The term in the denominator 4 (***l*** *·* ***n***) (***n*** *·* ***v***) comes from the derivation of the microfacet model and is usually included in most physically plausible models. *D* is the Normal Distribution Function (NDF), *F* is the *Fresnel* reflection coefficient (Eq. 4), and *G* is the Geometry shadowing and obstruction function (GSF). The NDF (*D*) statistically approximates the proportionate surface area of microfacets precisely aligned to the *halfway vector* (*h*). Additionally, it determines the highlight’s size and shape (*isotropic* or *anisotropic*). Several NDFs approximate the previously mentioned alignment of the microfacets, given a *roughness* parameter. Horizon uses NDFs based on the Generalized *Trowbridge-Reitz* (GTR) equation (Trowbridge and Reitz, 1975):

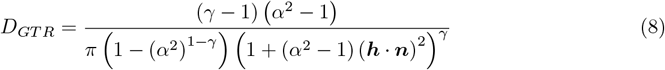

Where *α* = roughness^2^ is a remapping of the *roughness* parameter that improves the perception of shiny materials, and *γ* is a value greater than 0. Horizon includes two fixed specular lobes using GTR. The first and main lobe uses *γ* = 2, and the second lobe uses *γ* = 1. The main lobe describes the base material and may be *anisotropic* and/or *metallic*. The second lobe represents a *clear coat* layer over the base material and is thus always isotropic and non-metallic. It is important to notice that for *γ* = 1, there is a singularity at that point. Therefore, we need to take the limit as *γ →* 1, which produces this alternate form:

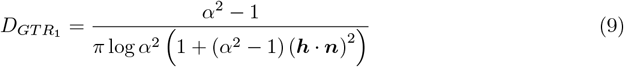

On the other hand, for a value of *γ* = 2, GTR is equivalent to the Ground Glass Unknown (GGX) model (Walter et al., 2007; Heitz, 2018):

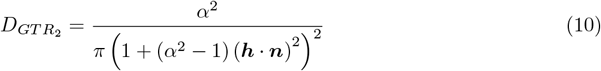

As mentioned before, Horizon’s main specular lobe could be *anisotropic*, which means that *roughness* distribution occurs unevenly in two directions. In most cases, these directions are the *tangent* and the *binormal* of a point on the surface. In such cases, the GTR takes the following form:

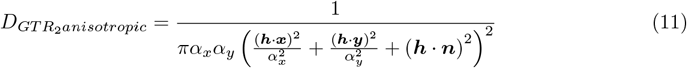

Where, *α*_*x*_ = roughness^2^*/ar* and *α*_*y*_ = roughness^2^ *\* ar* represent the *roughness* variation in two directions (*x* and *y* respectively). The *ar* parameter refers to an aspect ratio between the two directions, it depends of a single parameter (*anisotropic*), and is calculated as follows: 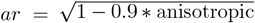, where the 0.9 factor limits the aspect ratio to 10 : 1.

Finally, the GSF (*G*) statistically approximates the relative surface area of lit and visible microfacets aligned to the *halfway vector* (***h***). In other words, the shadowing function accounts for not occluded, shadowed, or masked microfacets given a *view* and *light* directions. Similar to the NDF, the GSF takes a material’s *roughness* parameter as input, in which smoother surfaces have a lesser probability of occluded microfacets. The GSF used in Horizon is the separable *Smith* GGX derived by (Walter et al., 2007):

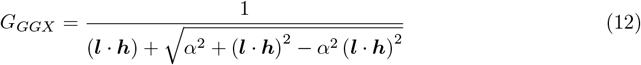

Which, in its anisotropic form, takes the following function:

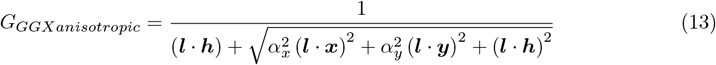

In summary, Horizon is fully compatible with Disney’s “principled” BRDF and adheres to its usability and interactivity principles. All the parameters listed here are normalized between 0 and 1 and behave similarly as indicated in (Burley and Studios, 2012). Concisely, we support the following material parameters: *Subsurface*, which controls the bi-layer blending between a surface’s outer and inner layers, from 0 to 1, soft to solid blending. *Metallic* regulates the conductive property of the material, where 0 is used for dielectric surfaces and 1 is used for metallic surfaces. *Specular* specifies the amount of incident reflection, being 0 a low intensity and 1 a high intensity specular light. *Specular Tint* interpolates the color of the incident light between an achromatic specular reflection (0) in white and the surface’s base color (1). *Roughness* controls how smooth the surface should be, in which 0 is used to represent smooth surfaces, and 1 is used to represent rough surfaces.

*Anisotropic* determines the shape of the specular reflections, where 0 creates isotropic reflections, and 1 anisotropic specular lights. *Sheen* models specific edge reflections on uneven surfaces, in this parameter values from 0 to 1, represent low to high intensities of such specular highlights on the edges. *Sheen Tint* is similar to *specular tint* with the exception that this parameter only affects the specular reflections determined by the *sheen* parameter. *Clear Coat* describes a second specular lobe added on top of the original surface. Here, the values between 0 to 1 define the intensity of the second layer. Finally, *Clear Coat Gloss* sets the smoothness of the second specular lobe created by the clear coat parameter. From 0 to 1, a mate to a satin effect of the clear coat lobe.

An additional parameter is provided as an independent shader. This shading pipeline is the first attempt to bring refractive representations to Horizon. In this case, we provide support to building translucent materials such as glass and liquids with light *absorption*. In order to achieve such effect, we use the *Beer-Lamber* law which is given by the following expression:

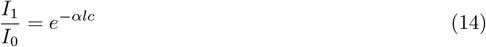

Where *I* represents light intensity, being *I*_0_ the intensity of the incident light, *I*_1_ the intensity of light that passes through the material, and the fraction represents the absorption rate. *α* is the absorption coefficient of the material. *l* is the distance the light travels to traverse the object. Finally, *c* is the concentration of absorbing elements within the material. Fig. 3 illustrates different levels of *absorption* with light information extracted from Image Based Lighting (IBL) textures.

**Figure 3.**
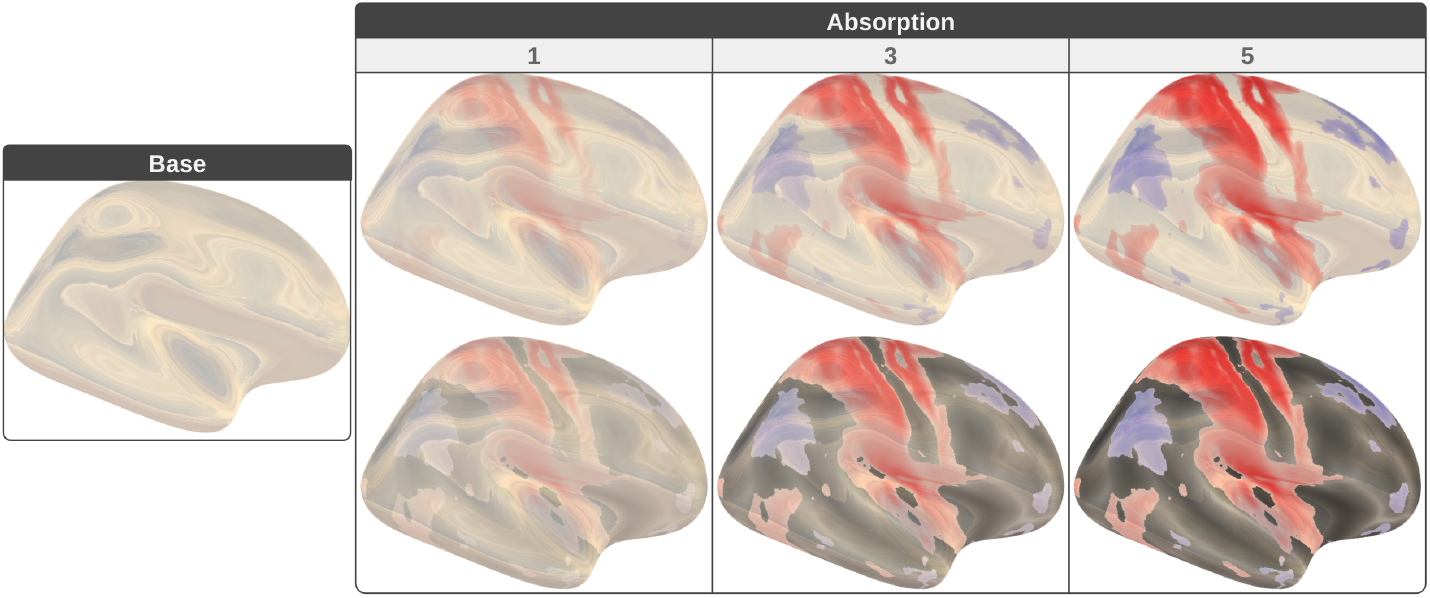
Gallery of physically-based glass-like shading with *absorption* parameter. **Base** contains a glass-like surface with reflected and refracted colors extracted using Image Based Lighting (IBL). Then, three different levels of ***absorption*** (1, 3, and 5) are used to combine the colors from the IBL with a statistical map (**top**) and with interpolated colors from the anatomical structure of the brain (**bottom**).

### 2.2. Accessing advanced machine learning data structures

Many medical imaging methods generate big data sets that need to be displayed. One of these techniques is fiber tracking algorithms which generate millions of streamlines (Rheault et al., 2015; Smith et al., 2015). For real-time applications, finding new ways to handle and simplify these large data sets are now a necessity. For that reason, Horizon provides easy ways to visualize such datasets’ matrices and time series. More importantly, it provides a mechanism to access hierarchical data structures straightforwardly and interactively. For example, Horizon can swiftly access the dynamically generated tree of clusters built by QuickBundlesX (QBX). QBX is an advanced, unsupervised machine learning algorithm that substantially improves over its predecessor QuickBundles (QB) (Garyfallidis et al., 2012). QuickBundles is one of the most efficient algorithms for clustering streamlines using streamline distances (Reichenbach et al., 2015), with a complexity of *O*(*kN*). *k* is the number of clusters, and *N* is the number of streamlines. Because *k* is usually much smaller than *N*, the algorithm’s complexity is near *O*(*N*) with large distance thresholds (e.g., 30mm). However, when small thresholds (e.g., 10mm) are used, the number of clusters increases considerably, and the complexity escalates accordingly. Additionally, it is essential to highlight that when *N* increases, so do *k*, i.e., the more streamlines in the dataset, the more clusters will be created. QBX is an online method that allows the clustering of large amounts of streamlines sequentially and in multiple resolutions by only passing one time through the whole streamlines. QBX does this by building a tree of centroid nodes on the fly, each layer representing a different distance threshold. As in QB, the distance metric used to compare the streamlines with the centroids of the clusters is the minimum average direct and flip distance (MDF) (Garyfallidis et al., 2012).

In detail, for a set of streamlines and a set of distances (e.g., 30mm, 25mm, 20mm), the algorithm will create a tree with as many levels as the number of input distances in descending order (Fig. 4). Then, each streamline will traverse the tree from the root to the leaves in the following way: At each level, the MDFs between the streamline and that level’s centroids are calculated and evaluated sequentially. During this process, the streamline is assigned to that cluster if the MDF is lower or equal to that level’s distance threshold. Otherwise, if the streamline is not within the radii of one of the existing centroids, then that streamline becomes a centroid at that tree level. This process is repeated at the following distance level until the last, and most minor distance threshold is evaluated. In other words, QBX dynamically creates a tree in which new centroids are added when the closest node to a streamline is far enough (MDF is greater than the threshold of the current layer) from the sibling nodes of that level (Fig. 4). Otherwise, the centroid of the closest node is updated, and then the process is repeated for its child nodes.

**Figure 4.**
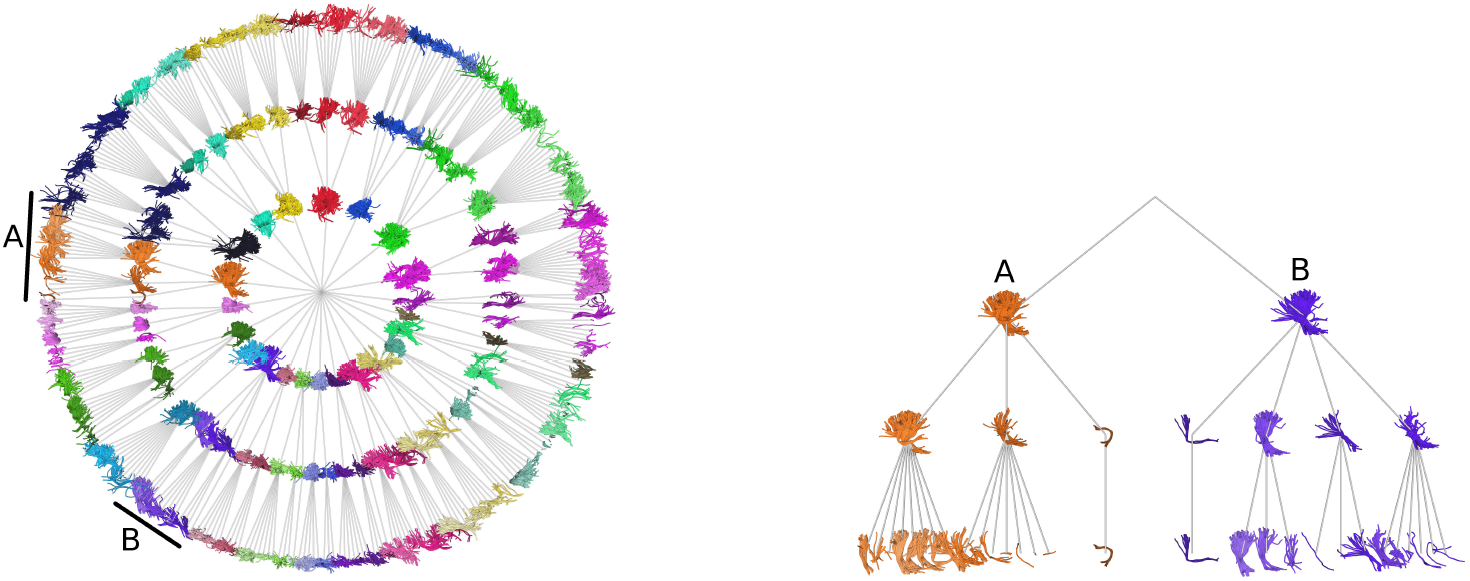
Example of a tree obtained from QuickBundlesX (QBX). On the **left**, the entire tree of centroids for a whole brain tractogram. For clarity, only the distance levels 30mm, 25mm, and 20mm are shown. On the **right**, a close-up of the subtrees **A** (orange) and **B** (purple) represent the left and right Corticospinal Tract, respectively.

This method returns a valuable tree of centroids at different distance resolutions. This structure allows to query streamlines and efficiently processing subsets of them by looking only at their vicinity. Computing the relative position of a streamline in a tractogram is generally an expensive operation because it involves a pairwise operation with each streamline in the dataset. However, this tree offers the possibility of quickly and efficiently knowing streamline’s surroundings. As in many tree traversal methods, the process starts at the root and recursively goes through the relevant child nodes until the leaves are reached. The criterion for finding the relevant centroids is based on the MDF, particularly the minimum distance. Although this new tree offers a better search structure, that is not its only application. Horizon takes advantage of this tree to speed up the visualization of large sets of streamlines, as can be seen in Fig. 5. In addition, the tool becomes more responsive by visualizing only the centroids from the upper level, as illustrated in Fig. 5B. Displayed tubes are width encoded, i.e., wider tubes represent more populated clusters. The tree structure can also be quickly accessed to explore and interact with some clusters, as shown in the expanded centroid in Fig. 5C. Nevertheless, this interaction is not limited to expanding the tree, but it also allows hiding unnecessary information so that neuroscientists can focus on exploring their bundles of interest, as can be seen in Fig. 5D. Furthermore, due to the hierarchical structure of the tree, this process can be repeated until the desired bundle is segmented, see Fig. 5E. Horizon uses custom shaders to achieve such a level of real-time responsiveness. For example, every time a centroid is selected, the color change is computed only for that centroid in the GPU by a fragment shader.

**Figure 5.**
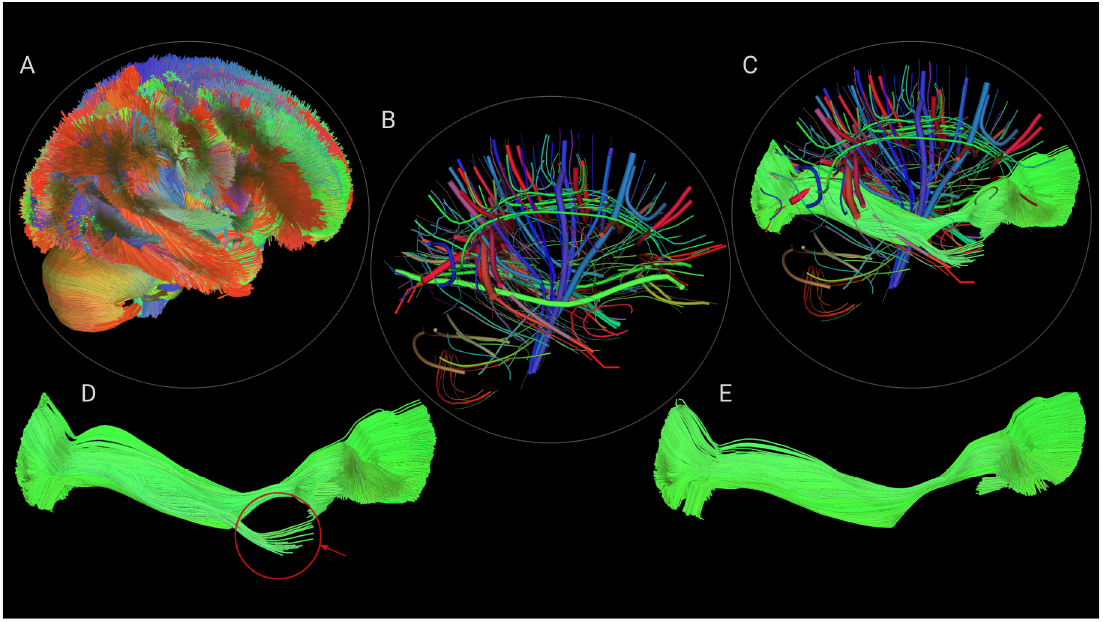
Horizon allows interaction with labeled tractograms directly. For example, a user starts by loading a full brain tractogram (**A**) which gets automatically clustered into bundles (centroids shown in **B**). Then, the user can expand any selected centroid (shown in **C**) or decide to hide the other clusters/centroids (shown in **D**). That process (cluster/selection/hide) can be repeated multiple times to remove subclusters (see red arrow) to obtain refined results, as shown in **E**.

### 2.3. Data

3T dMRI acquisition of subject 100307 from the Human Connectome Project (HCP) (Van Essen et al., 2013) was used. This dMRI acquisition (Sotiropoulos et al., 2013) has 1.25*mm* isotropic resolution, with 3 *b*-values (1000, 2000, 3000*s/mm*^2^) and a total of 270 gradient directions (90 per shell) and 18 *b* = 0 images. Voxelwise fiber orientation distribution functions (fODFs) of order 8 were reconstructed with DIPY. Probabilistic partial volume estimation (PVE) maps with FSL fast (Zhang et al., 2010) to extract the white-matter/gray-matter (wm/gm) interface and include/exclusion probabilistic regions. From the wm/gm interface, a seeding mask was created with a density of 10 seeds per voxel. Next, the inclusion and exclusion masks were used to set up an anatomically constrained stopping criterion. Then the tractogram was generated using the particle filtering probabilistic tracking algorithm (Girard et al., 2014) with its default parameters.

## 3. Results

### 3.1. Sheen as a new way to represent transparency

Horizon’s integrated PBR engine allows the exploration of different combinations of the “principled” parameters to highlight visualization features. In Fig. 6, the parameters *sheen* and *sheen tint* have been used to highlight the outline of a cortical surface displaying 14 regions of Destrieux’s atlas (Destrieux et al., 2010). Fig. 6A, shows the selected regions projected on FreeSurfer’s average cortical surface (Fischl et al., 1999). Then we computed a functional connectivity network (FCN) from a subject of (Nooner et al., 2012). The FCN was placed in the scene and used to test two different transparency approaches. In Fig. 6B, the surface’s opacity is gradually reduced until a good visual trade-off is achieved between the surface and the network. On the other hand, in Fig. 6C, *sheen* captures foldings and crevices on the surfaces and increases the specular reflection of those areas, which is perfect for outlining the sulci and gyri of the brain cortex. Seizing this property is possible to use *sheen* as a form of transparency that does not require changes in the opacity of an object but still retains depth information, as can be seen in the marked areas of Fig. 6C. It is also worth mentioning the property *sheen tint* because it gives control over the colors of the sheen’s specular reflections. This usage of *sheen* and *sheen tint* does not negatively impact or compromise the application’s performance.

**Figure 6.**
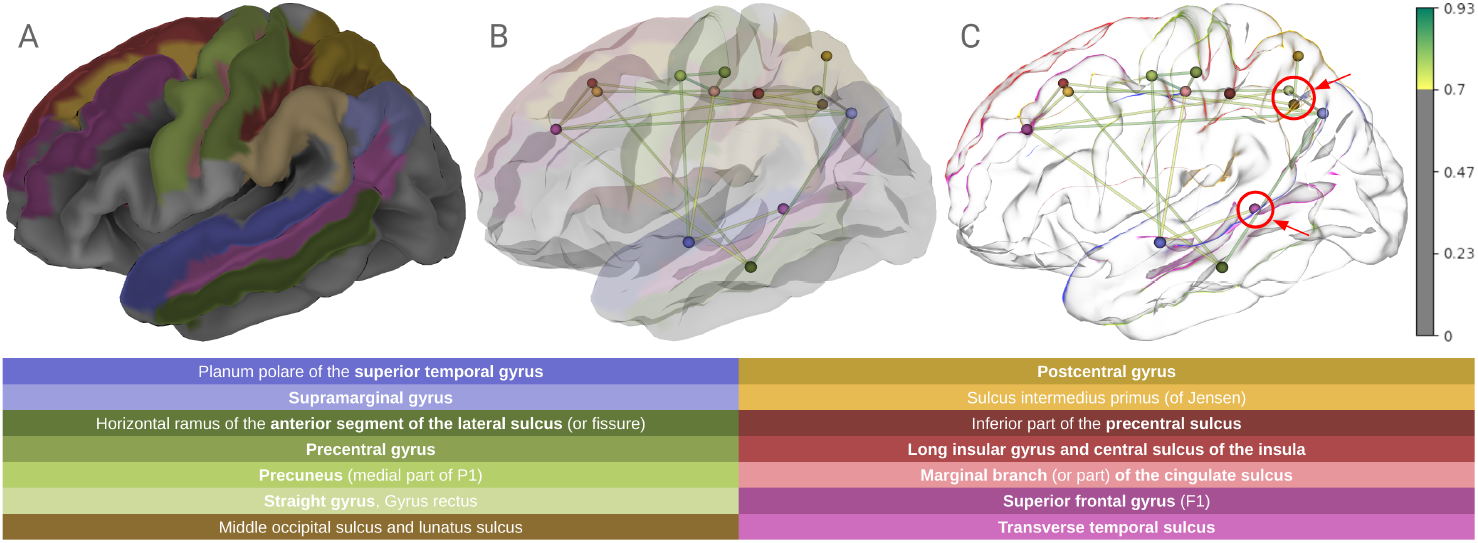
Visualization customization using physically-based shading. E.g., In **A**, atlas regions are projected on a cortical surface. **B** and **C** display a connectivity network alongside the cortical surface. In **B**, the surface opacity is reduced until the graph becomes visible. While in **C**, *sheen* highlights the surface’s silhouette without compromising the opacity. This effect preserves depth information, as seen in the marked regions (red circles).

### 3.2. Enhanced line and streamtube shading

Most existing tools for visualizing fibers render them as unshaded lines or polygonal tubes. Unfortunately, the use of unshaded lines gives no cues about the shape of the fibers, as shown in Fig. 7. On the other hand, polygonal tubes require many polygons to achieve high image quality. Unfortunately, this might compromise rendering performance. Additionally, neither of these methods communicates the coherent structure of a collective of fibers.

**Figure 7.**
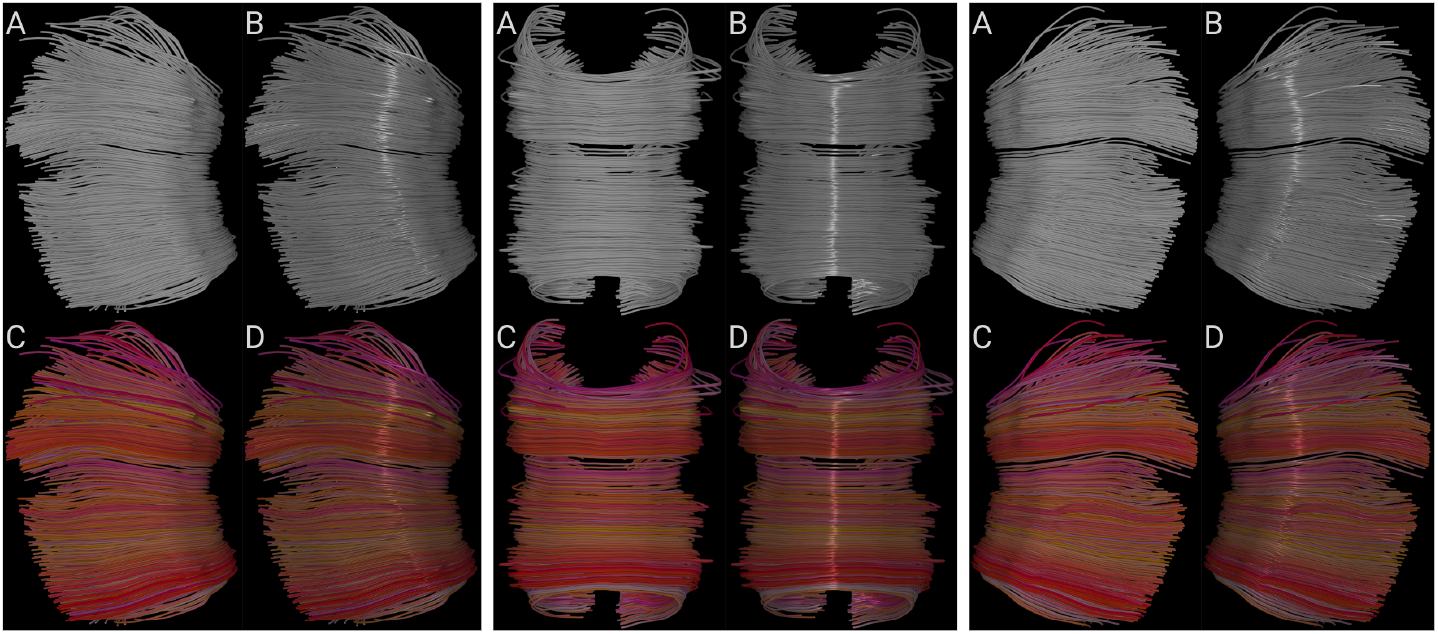
Comparison of lighting and coloring techniques on three views of the midsection of the Corpus Callosum. In each view, bundle with a plain gray color (**A**). Bundle with a plain gray color and *anisotropic* lighting (**B**). Bundle with colors interpolated from each fiber direction (**C**). And bundle with interpolated colors and *anisotropic* lighting (**D**).

Visualization of fibers can be compared to human hair rendering since both aim to display many polyline-like objects with particular shapes and coherencies. These shapes and coherencies are essential for the user because they contain vital information about the dataset’s structure. In human hair rendering, they represent the hairstyle that the viewer then interprets. In medical imaging visualization, they represent the underlying anatomy associated with its biological process. Hair rendering techniques use illumination as part of the identity of each hairstyle since it helps to explain shape in a better way. Similarly, medical imaging visualization could use illuminated lines to perceive the form of entire bundles in a better way (Stalling et al., 1997; Mallo et al., 2005). Horizon’s *anisotropic* parameter could be used to illuminate lines and streamtubes, giving better and far more intuitive cues about the shapes of the fibers than flat shading, even if color coding of the local fiber orientation is used (see Fig. 7).

### 3.3. Photorealistic stained glass shading

Photorealistic translucent surfaces, when appropriately rendered, provide a more natural see-through effect. This effect is ideal for conditions where extra information lies inside or beneath another surface of relevance. Additionally, stained glass representations or, in Horizon’s case, refractive medium with *absorption* will eventually allow for highlighting spatial information on the outer surface, such as heatmaps (Fig. 8) while still providing a fully translucent element. In Fig. 8, the *absorption* will allow scientists to enable or disable such heatmaps without reducing the opacity of the external surface.

**Figure 8.**
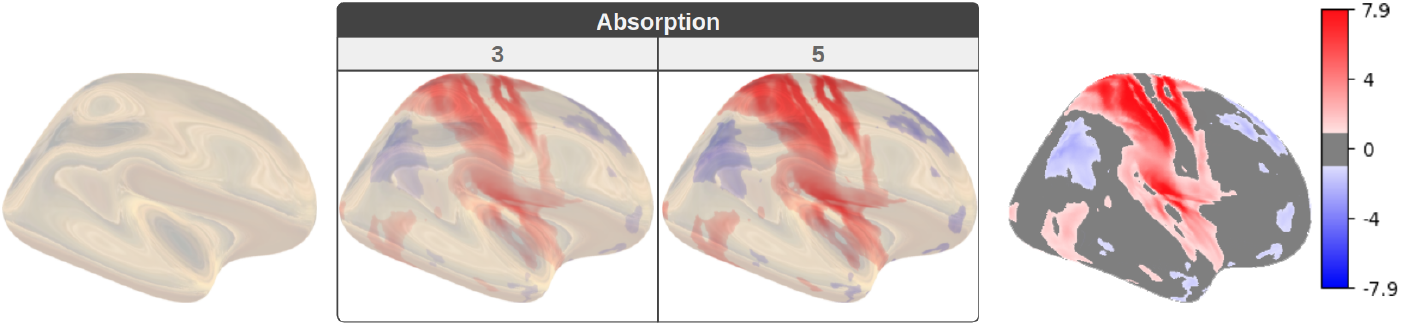
Comparison of fMRI statistical maps. A cortical surface with Horizon’s glass shader and IBL (**left**). Then, two different levels of *absorption* (3 and 5) (**center**) are used to combine the colors from the IBL with a statistical map. Last, the same statistical maps and interpolated colors on non-glass surfaces (**right**).

### 3.4. Using QBX to improve streamlines visualization

To demonstrate the improvements included in QBX over its predecessor, we ran probabilistic tracking using anatomical priors and randomized seeds generation in the white matter on a subject from the HCP. This configuration generated from 1 million to 5 million whole brain tractograms. Since QBX requires additional parameters compared to QB, the distances used in this experiment were the following 30mm, 25mm, 20mm, and 15mm, which produced a tree with four layers giving access to the clusters of those resolutions. Compared to the single-level clusters of QB.

Then the execution time for QB and QBX was compared and illustrated in Fig. 9. QBX exhibited a speedup of 22X to 25X over QB, meaning that 5 million streamlines can be grouped in less than 5 minutes on hardware equivalent to a standard laptop (Intel core i7, 1.8 GHz). In contrast, QB needed at least 2 hours to perform the same task. These performance improvements allow for an interactive experience, as shown in Fig. 5C. In which a centroid has been expanded, displaying the streamlines that belong to that cluster node. Then, the tree is used to refine the displayed bundle by discarding undesired streamlines (Fig. 5D and 5E). It is worth mentioning that, in this case, Horizon directly accesses the buffer of clusters and moves back and forth from the centroid to the cluster representation. Users can click on a centroid or cluster and move from one reproduction to another. Tasks like this are becoming more popular to improve data exploration.

**Figure 9.**
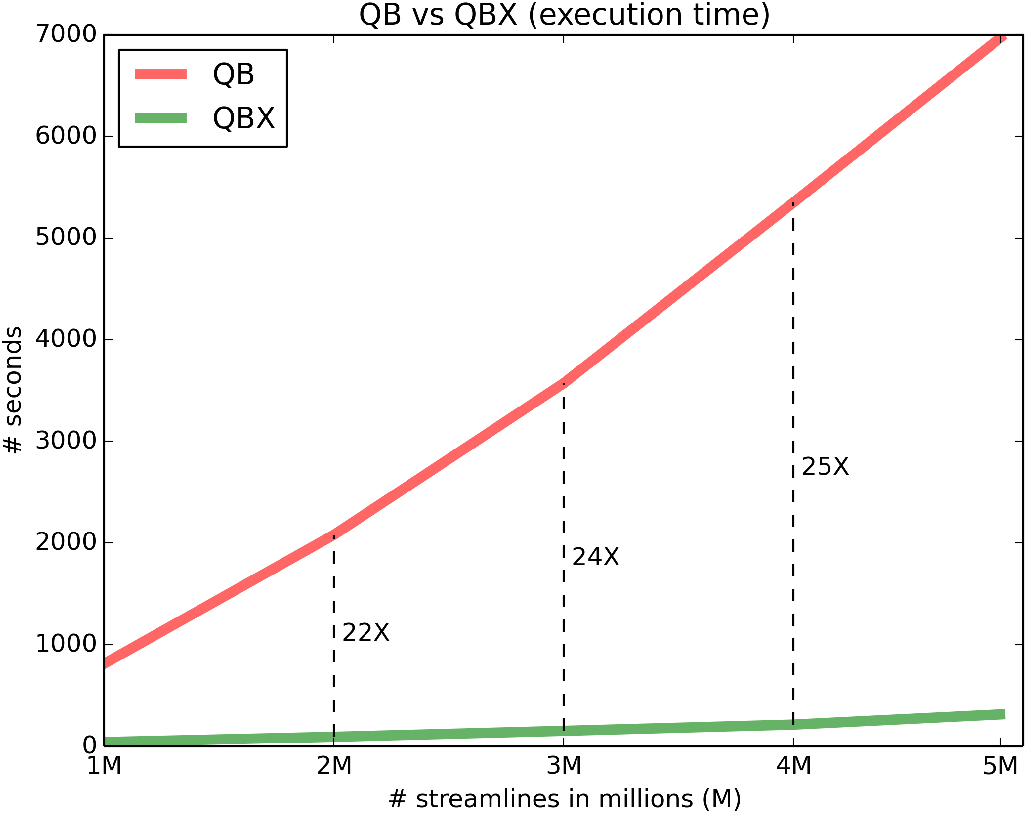
Performance comparison between QuickBundles (QB) (Garyfallidis et al., 2012) and QuickBundlesX (QBX). Lower execution times mean faster clustering time. QBX is about 20X faster than QB, which was already a fast method for clustering streamlines.

## 4. Discussion

Horizon is an advanced rendering engine and software for medical imaging visualization. It provides high efficiency by exploiting parallel programming in the GPU and offers high flexibility via its API interface in Python. The main focus of this paper is on physically-based rendering capabilities, which are offered in Horizon using shaders both for reflection and refraction. The proposed rendering engine shares some of the fundamentals of the famous Disney BRDF explorer from which it is inspired. Its parameters are easy to interpret and use without compromising realistic results but still give users the flexibility to create surreal visualizations. Additionally, the light simulations were validated using the Material Exchange and Research Library (MERL) database (Matusik, 2003), which provides a degree of certainty in the case our users would want to create highly accurate photorealistic visualizations.

In this introductory paper, we use the new rendering engine to explore how it can help scientists to get more insights from their data. The included parameters allow for novel representations of the data. For example: a) we demonstrated how sheen reflections could be used as an alternative to traditional opacity and similar occlusion techniques. This idea could be relevant for scientists working with cortical surfaces in conjunction with connectivity networks or tractography-based approaches. b) We demonstrated how anisotropic illumination, a technique borrowed from hair rendering, can improve the appearance of elongated objects, such as streamlines and streamtubes. c) We used a novel refractive shader to explore color blending intensities of heatmaps with glass-like rendering techniques. This shader sets the basis to explore different Bidirectional Distribution Functions (BDF) and effects like Screen Space Refractions. Finally, d) we showed how combining machine learning (ML) algorithms and data structures with shaders can accelerate the visualization of complex elements such as tractograms. These kinds of strategies are common in production rendering in which the size of the assets and associated information is considerably large, so efficient ways to handle and visualize the data constitute the main challenge. In summary, Horizon adds multiple new dimensions for investigating the data. Every parameter (or their combinations) of the BRDF can be used to create a new story about the geometry and the features of medical datasets. Advanced shading effects have been explored, and some efforts have found great value in ambient occlusion and shadowing (hard and soft) techniques. Therefore, adding such extensions to our rendering engine would be appropriate as the first next step. Additionally, a natural extension of this work would be transitioning from a BRDF to a bidirectional scattering distribution function (BSDF) with subsurface scattering capabilities. Lastly, when combined with ray tracing-based rendering approaches, the included lighting methods can significantly improve the overall perception of the generated images. One of the many benefits we consider relevant for scientific visualization is indirect or global illumination techniques. However, Horizon is currently only supporting ray-marching. We left ray tracing and global illumination as future work because these methods are now too slow for real-time application in consumer hardware (general-purpose desktops and laptops that most scientists use).

A noticeable impact of this work is that Horizon brings some of the latest rendering technology to the medical imaging domain and offers it in a way that adheres to open standards. Therefore, Horizon creates a much-needed environment where multiple domains can meet, build and extend. The provided API is fully customizable and can be modified to include other (non-PBR) lighting techniques. Developers can change or edit any internal parts of the code and create new products without licensing restrictions.

### 4.1. Ecosystem and availability

Horizon is built to allow the user to interact with multiple objects (streamlines, surfaces, volumes, glyphs, or assemblies of those) and user interface components using the same API, as shown in Fig. 10. Furthermore, this API allows to a) directly program shaders on different visual actors, b) interact with and pick any object, c) build scripted animations with timelines, d) work seamlessly with DIPY, NiBabel, matplotlib, numpy, scipy, VTK, TensorFlow and other libraries of the pythonic ecosystem, e) allow web visualization, desktop and VR visualization (see Fig. 10) with no changes in the code, f) visualize multiple subject data at the same time in grid or layered views, g) load data from the command line, the python script, the web or directly from the window, h) interact between clusters, bases (spherical harmonics) and final data. Horizon is provided together with DIPY. The main corpus of shaders is developed in the FURY library (Garyfallidis et al., 2021), which is one of DIPY’s visualization dependencies.

**Figure 10.**
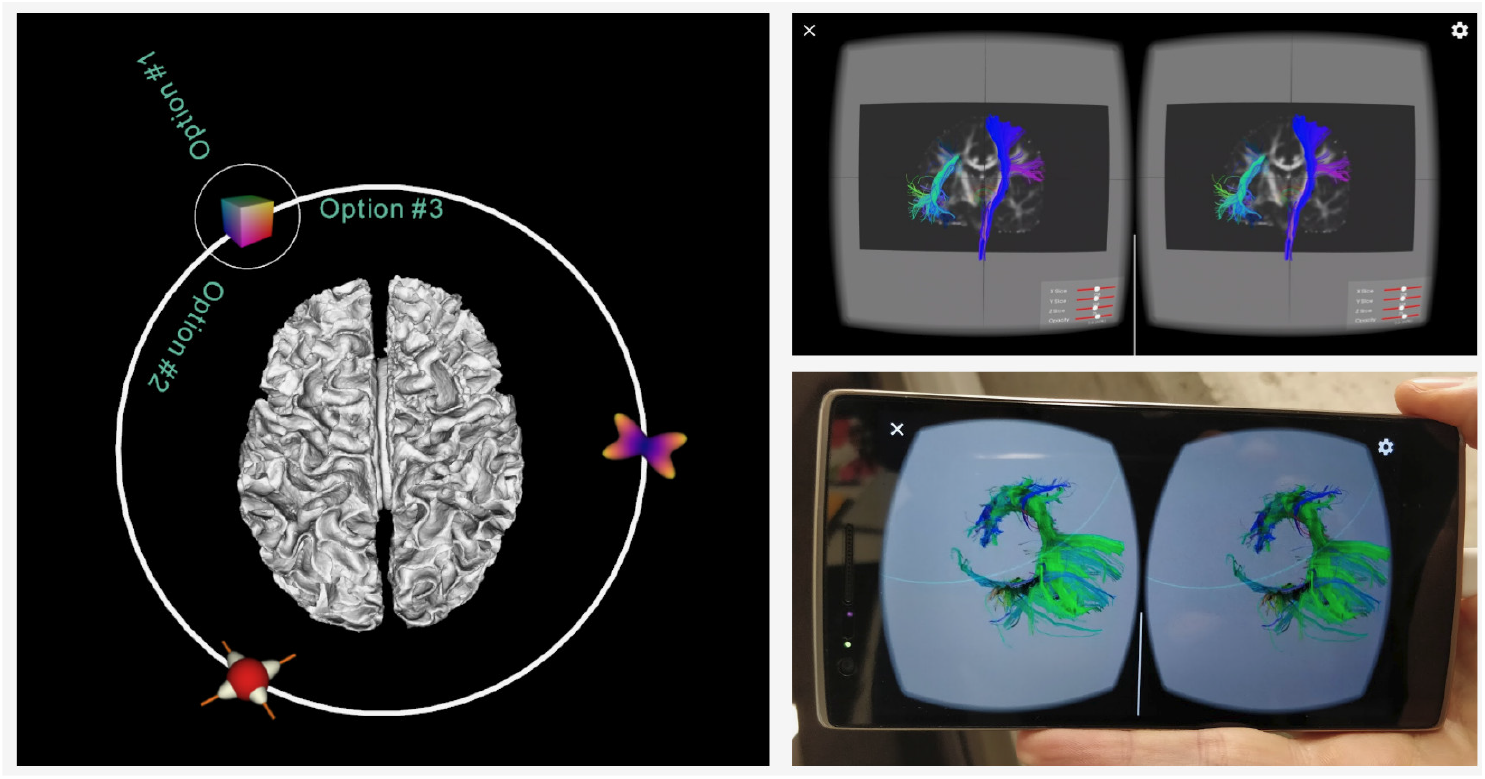
This figure demonstrates two advanced interfaces. On the left is an orbital menu that can be attached to any 3D object. On the right is a virtual reality application that uses OpenVR integration to render stereo image pairs on a mobile phone for use in a VR headset.

## 5. Conclusion

We presented Horizon, a new software engine to democratize advanced visualization for medical imaging. Horizon brings state-of-the-art shading techniques commonly seen in the gaming and animation industries. It presents them in an easy-to-use and understandable way so medical imagers can focus on visualizing their data rather than studying complex light transport concepts. In this way, Horizon enables and accelerates discovery by facilitating access to technologies usually out of reach for most medical imagers. Horizon comes with innovations, such as using sheen for transparency, photorealistic glass animations, illuminated tractography with anisotropic shaders, and provides shader access to advanced ML algorithms and data structures in real-time. For the latter, an example for accessing hierarchical clusters of tractograms was provided. Horizon is publicly available with DIPY.

## Data Availability

All data produced in the present study are available upon reasonable request to the authors.

## 6. Acknowledgements

This work was conducted in part using the resources of the Technology for Research division of the University Information Technology Services (UITS) at Indiana University, Bloomington, IN. This work was supported by the National Institute Of Biomedical Imaging And Bioengineering (NIBIB) of the National Institutes of Health (NIH) under Award Numbers R01EB027585 and 2R01EB017230-05A1. FURY is partly funded through NSF 1720625 Network for Computational Nanotechnology - Engineered nanoBIO Node (Klimeck et al., 2008).

### 7. Appendix

QBX can dramatically reduce execution time for grouping streamlines at low distance thresholds and building hierarchies as shown in Fig. 9.

Recent technologies, such as virtual reality (VR), are fully compatible with Horizon, thanks to its flexible API, as shown in Fig. 10.

